# Intracranial approach for sub-second monitoring of neurotransmitters during DBS electrode implantation does not increase infection rate

**DOI:** 10.1101/2021.11.16.21266363

**Authors:** Brittany Liebenow, Michelle Williams, Thomas Wilson, Ihtsham ul Haq, Mustafa S. Siddiqui, Adrian W. Laxton, Stephen B. Tatter, Kenneth T. Kishida

## Abstract

**Introduction:** Currently, sub-second monitoring of neurotransmitter release in humans can only be performed during standard of care invasive procedures like DBS electrode implantation. The procedure requires acute insertion of a research probe and additional time in surgery, which may increase infection risk. We sought to determine the impact of our research procedure, particularly the extended time in surgery, on infection risk.

**Methods:** We screened 607 DBS electrode implantation procedures performed at Wake Forest Baptist Medical Center between January 2011 through October 2020 using International Classification of Diseases (ICD) codes for infection. During this period, 116 cases included an IRB approved 30-minute research protocol, during the DBS electrode implantation surgery, to monitor sub-second neurotransmitter release. We used Fisher’s Exact test (FET) to determine if there was a significant change in the infection rate following DBS electrode implantation procedures that included, versus those that did not include, the neurotransmitter monitoring research protocol.

**Results:** Within 30-days following DBS electrode implantation, infection was observed in 7 (1.43%) out of 491 procedures that did not include the research procedure and 2 (1.72%) of the 116 procedures that did include the research procedure. Total infection rates (i.e., not constrained by 30-day time window) for all non-research cases was 28/491 (5.70%) and only 4/116 (3.45%) for research inclusive cases. Notably, all types of infection observed were typical of those expected for DBS electrode implantation.

**Conclusion:** Total infection rates are not statistically different in patients who performed the research procedure (3.45% vs. 5.70%; p = 0.4872, FET) and not statistically different across research and non-research groups within 30-days following the research procedure (1.72% vs. 1.43%; p = 0.684, FET). Our results demonstrate that the research procedures used for sub-second monitoring of neurotransmitter release in humans can be performed without increasing the rate of infection.

## Introduction

There is great promise in leveraging opportunities in the operating room (OR) to conduct human neuroscience research. Deep Brain Stimulation (DBS) in particular lends itself to research, as the procedure typically entails intraoperative electrophysiological assessments of neural targets; thus, research data can be acquired with relatively minor protocol changes [1–5]. This has allowed research teams to make breakthrough discoveries using data collected in the OR during DBS electrode implantation procedures [1–16]. Notable innovations include first-of their kind measurements of neurotransmitters [1–5,7,8], including dopamine [1,2,4,5,12], serotonin [3,4,12], and adenosine [7,8] in humans. Other advances include single unit recordings from substantia nigra [6] and expanding DBS targeting to provide symptom relief in treatment-refractory depression, substance use disorder, Alzheimer’s disease, and obsessive-compulsive disorder [9–11,13–16].

One potential limiting factor in conducting translational research in the OR is the possibility that the added OR time necessary to conduct experiments may increase infection risk [17,18]. Infection risks following DBS surgeries are well described and provide a good basis for comparison. A metanalysis covering 1354 patients across 23 articles reported a 6.9% overall risk of infection following DBS electrode implantation surgeries [19]. Similarly, one large single-center study of 447 DBS patients identified an overall infection rate of 5.82% (26 patients); this study also identified a 30-day infection rate of 2.01% (9 patients) [17]. This 30-day infection rate is corroborated by other large, single-institution studies, including a study of 273 patients with a median time to infection of 1 month that reported an infection rate of 3.1% across procedures for primary DBS electrode placement [20]. To date, however, no study has investigated whether adding OR time due to a predefined research protocol increases infection risk after elective surgery.

Our group has over a decade of data and experience measuring neurotransmitters while patients complete behavior tasks during DBS electrode implantation surgeries [1–5,12]. These experiments have added a maximum of 30 minutes to the scheduled OR time. Thus, we have ample research and surgical records to retrospectively explore whether there are group differences in post-operative infection rates between patients receiving DBS who participate in research (‘research’) and who do not participate in research (‘non-research). Here, we compare the 30-day and time unconstrained post-operative infection rates of research (N=491) and non-research (N=116) groups at our institution to investigate whether these experiments increased infection risk.

## Materials and Methods

The Institutional Review Board at Wake Forest University Health Sciences approved all procedures described for this retrospective study (IRB00064371) and for our ongoing research protocol in the DBS operating room (IRB00017138). Under IRB00017138, research participants gave their written informed consent prior to participation in the research study. The data reported in this manuscript are not publicly available due to containing identifying information that would compromise the privacy of patients and research participants whose data are deidentified and summarized in this study.

### Clinical Data

We screened all DBS electrode implantation procedures performed at Wake Forest Baptist Medical Center between January 2011 through October 2020 using International Classification of Diseases (ICD) codes for infection. These ICD codes included: T85.731 (Infection and inflammatory reaction due to implanted electronic neurostimulator of brain); T85.734 (Infection and inflammatory reaction due to implanted electronic neurostimulator, generator), 61867 (First Electrode with microelectrode recording, typical), 61868 (Second Electrode on same side with recording, other side), 95983 (Intraoperative analysis / programming), 61885 (For single electrode), 61886 (For multiple electrodes), 61880 (Electrode removal / revision), 61888 (Generator removal / revision, use for attaching previously placed lead); Z45.42 (DBS Phase / Stage 3, generator change).

We further screened the total number of DBS procedures using the medical record number (MRN) of all patients who received DBS and participated in our research protocol (Tab. 1). The research and non-research groups were examined separately.

**Tab. 1.** Total numbers of patients who are consented for, and ultimately complete, a research protocol during deep brain stimulation (DBS) neurosurgery. This data starts in April 2012 and continues through December 2020.

| Year | Total Patients Consented | Total Patients Completed | Total Patients Consented Only | % Patients Completed | % Patients Consented Only |
| --- | --- | --- | --- | --- | --- |
| 2012 | 4 | 4 | 0 | 100% | 0% |
| 2013 | 23 | 17 | 6 | 74% | 26% |
| 2014 | 20 | 14 | 6 | 70% | 30% |
| 2015 | 9 | 7 | 2 | 78% | 22% |
| 2016 | 13 | 11 | 2 | 85% | 15% |
| 2017 | 14 | 10 | 4 | 71% | 29% |
| 2018 | 24 | 18 | 6 | 75% | 25% |
| 2019 | 23 | 15 | 8 | 65% | 35% |
| 2020 | 9 | 8 | 1 | 89% | 11% |

Our research protocol includes measuring neurotransmitters from a research microelectrode that is put in place prior to the Tungsten functional mapping microelectrode [1–4,12]. Our research targets (the caudate, putamen, and thalamus) are anatomically superior to therapeutic targets used in DBS (the STN, GPi, and thalamus), thus use of our research electrode is typically performed just prior to functional mapping with tungsten microelectrodes (provided by FHC inc.) [1–4,12]. While the research team measures neurotransmitter levels, participants complete behavioral tasks displayed on a computer monitor and input decisions using a standard gaming controller [1–4,12]. This research protocol is limited to 30 additional minutes maximum added to DBS surgery time and requires an informed consent process that takes place ahead of the surgery [1–4,12]. Research records were also screened to determine the number of patients consenting to versus completing research protocols during their DBS surgeries.

### Statistics

All statistical analyses were conducting using RStudio [21]. We used Fisher’s Exact test (FET) to compare overall and 30-day infection rates following DBS electrode implantation procedures across the cohorts that did and did not participate in our DBS research protocol.

## Results

We identified 607 DBS electrode implantation procedures performed at Wake Forest Baptist Medical Center between January 2011 through October 2020 using our specific ICD codes for infection (Fig. 1). Of those 607 cases, 491 procedures across 487 patients met criteria for DBS cases that did not include the research protocol (‘non-research’), and an additional 116 procedures across 116 patients met criteria for DBS cases that did include the research protocol (‘research’). For the non-research group, the average age was 70.65 +/- 12.64 years, with 36.46% female and 63.54% male. For the research group, the average age was 68.17 +/- 9.73 years, with 25.86% female and 74.14% male.

**Fig. 1.**
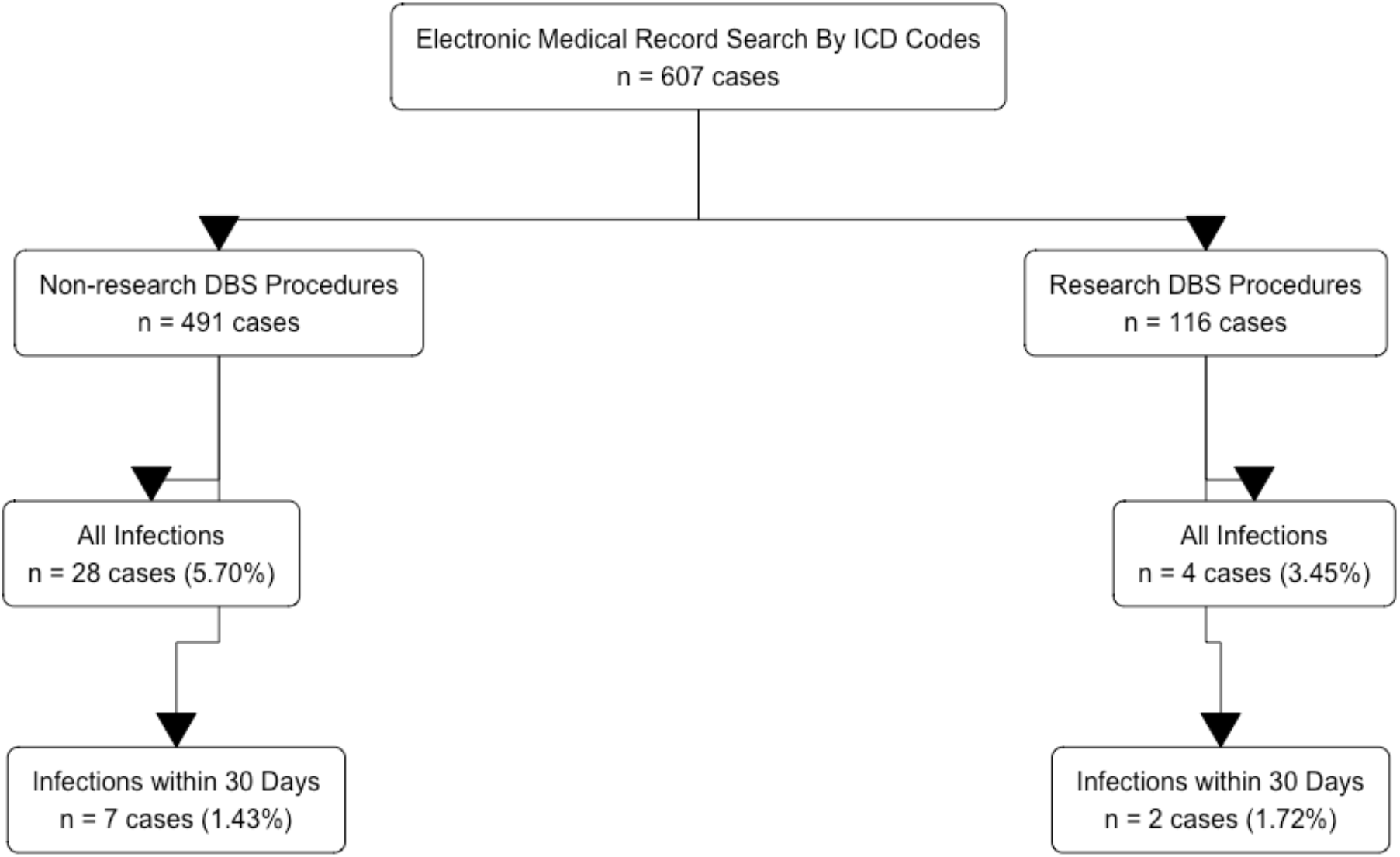
Flowchart demonstrating the acquisition and filtering of deep brain stimulation (DBS) procedures (or cases) from the electronic medical record (EMR) using International Classification of Diseases (ICD) codes.

Of the 491 non-research DBS cases, there were 7 infections within 30 days of the DBS procedure (1.43% of total cases) and 28 infections not constrained by 30-day time window (5.70%) (Fig. 2). Of the 116 research DBS cases, there were 2 within 30 days of the DBS procedure (1.72% of total) and 4 infections not constrained by 30-day time window (3.45%) (Fig. 2). Using FET, we determined that there is no statistically significant difference between the non-research and research DBS groups in the 30-day infection rates (1.43% vs. 1.72%; p = 0.684, FET), or the overall infection rates (5.70% vs. 3.45%; p = 0.4872, FET).

**Fig. 2.**
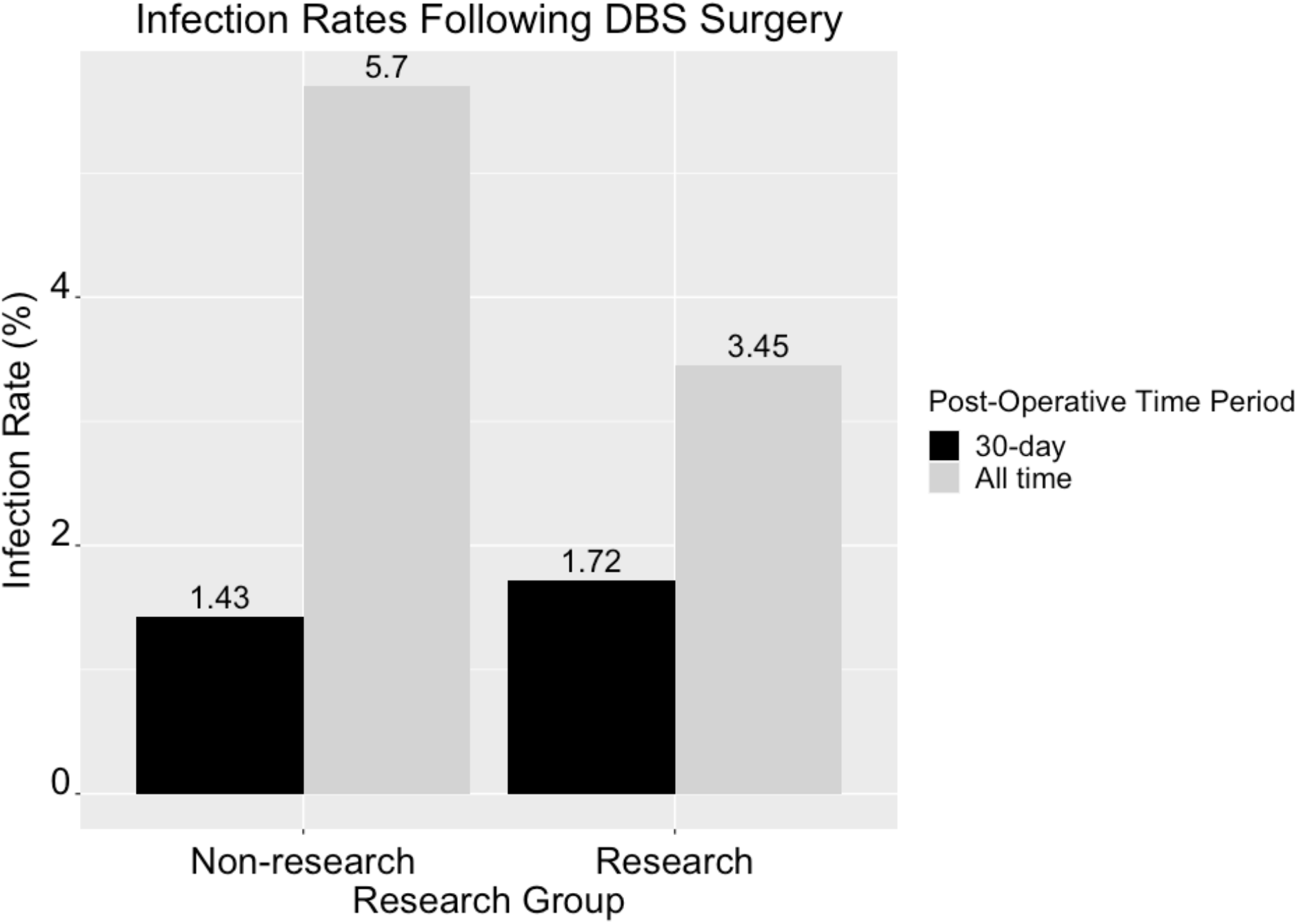
Comparison of infection rates across the research and non-research groups after deep brain stimulation (DBS) neurosurgery. Infection rates are also compared across the 30-day and unconstrained time windows after neurosurgery.

The infectious pathogens in the research group were reported to be: Staphylococcus aureus, Methicillin-sensitive Staphylococcus aureus (MSSA), Coagulase-negative staphylococci (CoNS), and Serratia marcescens. All of these pathogens have been reported in the literature as potential causes of post-operative infections after DBS, with the Staphylococcus genus the most common culprit [22].

We also identified the yearly number of patients who were approached for consent to participate in research, and the actual number of patients who completed the research study starting in April 2012 and continuing through December 2020 (Tab. 1). The diagnoses of consented patients were also identified (Tab. 2). Reasons for consenting but not completing a research protocol can be influenced by the individual anatomy of each participant (i.e. no good trajectory connecting striatum and target).

**Tab. 2.**
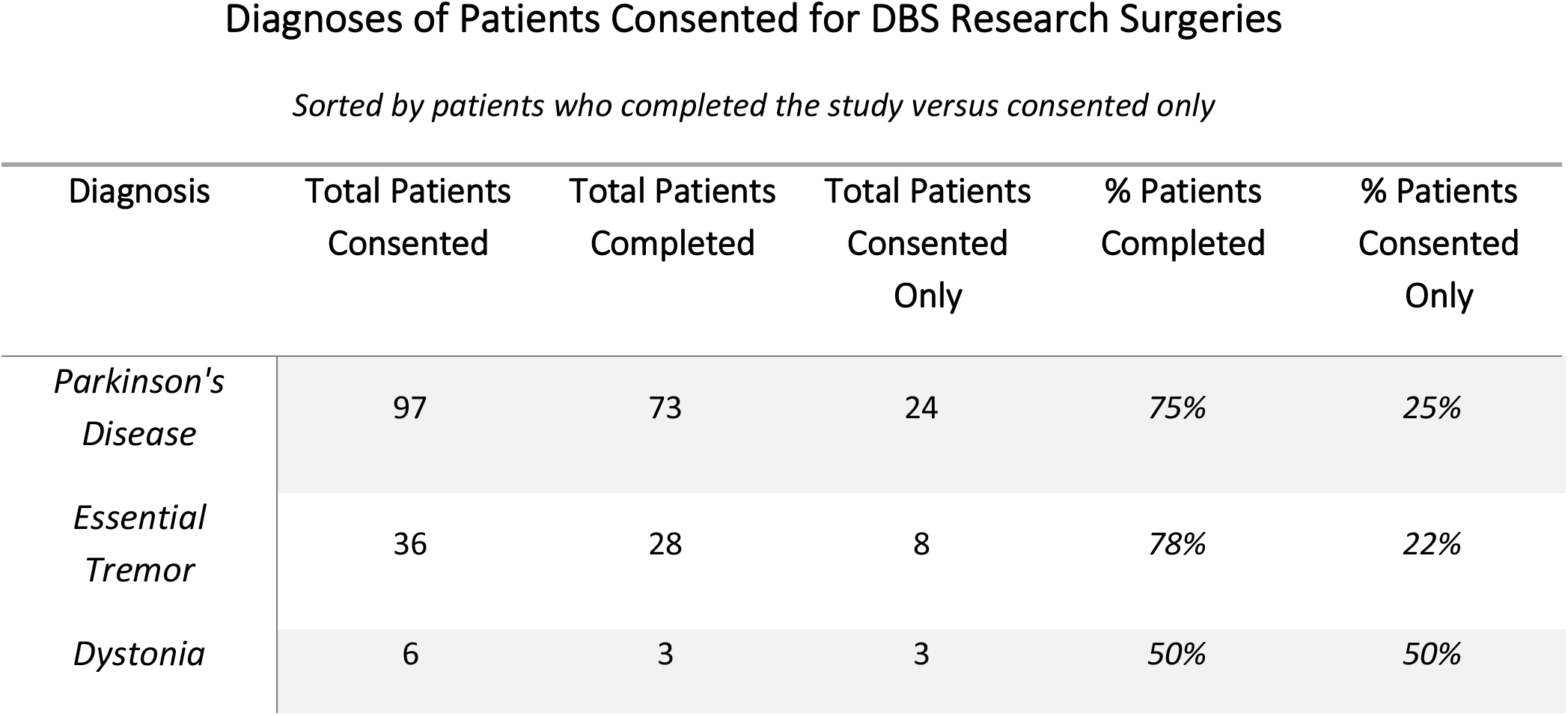
Diagnoses of patients who are consented for, and ultimately complete, a research protocol during deep brain stimulation (DBS) neurosurgery. This data starts in April 2012 and continues through December 2020.

| Diagnosis | Total Patients<br>Consented | Total Patients<br>Completed | Total Patients<br>Consented<br>Only | % Patients<br>Completed | % Patients<br>Consented<br>Only |
| --- | --- | --- | --- | --- | --- |
| *Parkinson's*<br>*Disease* | 97 | 73 | 24 | 75% | 25% |
| *Essential*<br>*Tremor* | 36 | 28 | 8 | 78% | 22% |
| *Dystonia* | 6 | 3 | 3 | 50% | 50% |

## Discussion/Conclusion

Our results investigating the infection rates of DBS surgery patients with and without a 30-minute invasive research procedure during DBS electrode implantation demonstrate that there is no difference in overall and 30-day infection rates between groups. This is consistent with the conclusion that these experiments can be performed without increasing the risk of infection in these patient populations. Further confidence in our results can be found in a comparison to studies reporting similar overall and 30-day infection rates to our results. Our 30-day infection rates of 1.43% for non-research and 1.72% for research procedures are comparable to—and even lower than— similar studies reporting 30-day infection rates of 2.01% [17] and 3.1% [20] where intracranial research was not performed. Likewise, our overall infection rates of 5.70% for non-research and 3.45% for research cases are comparable to, and lower than, similar studies reporting 6.9% [19] and 5.82% [17]. The infectious pathogens reported for the research infections (Staphylococcus aureus, MSSA, CoNS, and Serratia marcescens) have all been reported as potential infectious causes in post-operative DBS infections [22]. This provides further support that the 30-minute research period introduces no new infection related risks to patients.

There are a number of potentially influential factors in maintaining low infection rates while conducting translational research in the DBS OR. First and foremost, all research electrodes in our study go through independently validated sterilization procedures typical of all surgical equipment requiring sterilization [1–4]. During the research procedure, the neurosurgeon leads the clinical staff and maintains the sterile field while handling all equipment within the sterile field. We have limited the research protocol to a maximum of thirty minutes of additional time in surgery. Should the research tasks be delayed or extended for any reason the research activities are to be prematurely terminated at the thirty-minute threshold. This is done primarily to avoid unbounded delays in the surgery so that patient safety and comfort are maintained as much as possible. All patients who are candidates for DBS-electrode implantation surgery are offered the opportunity to participate. Those that choose to volunteer may be among the most likely to be capable of post-surgical selfcare that would aide in minimizing post-surgical infections. In this retrospective analysis, we do not have the appropriate data to assess this possibility, but it is potentially a major factor in the low infection rates we observe. The similarly low infection rates in the non-research group suggests that if this were the explanation for low infection rates then the comprehensive process of screening potential candidates for DBS-surgery at our institution would be the causative factor.

Research, including this current study, that shares information about the risks of observational human research studies during DBS are necessary to verify the anticipated safety of new applications of intracranial research protocols in the neurosurgery setting. The DBS-electrode implantation procedure is a safe and relatively low risk procedure as has been demonstrated repeatedly in the past [1–6]; it also affords unique access to areas of the human brain that have not been accessible in the past. Measurements of neurotransmitters, single units, and local field potentials in the human brain may provide new information about brain function and the mechanisms underlying disorders that may aid in improving the efficacy of DBS treatment [1–4,6,11–13] or in the development of novel neurosurgical goals. In addition to basic knowledge generation, these studies may also lead to the development of novel biologic markers of disease and treatment management. Our demonstration that these kinds of research protocols can be performed without an increase in infection rates should – with appropriate expertise, care, and consideration – encourage further intracranial investigation of human brain function. Our results show that intracranial recordings of sub-second neurotransmitter release in a time-extending research protocol utilizing a novel research probe are possible without increasing infection rates.

## Data Availability

The data reported in this manuscript are not publicly available due to containing identifying information that would compromise the privacy of patients and research participants whose data are deidentified and summarized in this study.

## Statement of Ethics

The Institutional Review Board at Wake Forest University Health Sciences approved all procedures described for this retrospective study (IRB00064371) and for our ongoing research protocol in the DBS operating room (IRB00017138). Under IRB00017138, research participants gave their written informed consent prior to participation in the research study. This statement is included in the manuscript.

## Author Contributions

Brittany Liebenow, Michelle Williams, Thomas Wilson, Ihtsham ul Haq, Mustafa S. Siddiqui, Adrian W. Laxton, Stephen B. Tatter, and Kenneth T. Kishida (all authors) have met all four requirements for authorship designated by the ICMJE, which include: substantial contributions to the conception or design of the work; or the acquisition, analysis, or interpretation of data for the work; drafting the work or revising it critically for important intellectual content; final approval of the version to be published; and, agreement to be accountable for all aspects of the work in ensuring that questions related to the accuracy or integrity of any part of the work are appropriately investigated and resolved.

